# The Vocabulary Test of the California Cognitive Assessment Battery (CCAB)

**DOI:** 10.64898/2026.10.25.26366434

**Authors:** David L. Woods, Mike Blank, Kathleen Hall, Isabella Jaramillo, Kristin Geraci, Peter Pebler

## Abstract

**Objective:** We report the design and metric properties of the vocabulary test of the California Cognitive Assessment Battery (CCAB), which adaptively delivers 24 trials from a 600-word list ordered by difficulty, and describe the demographic variables associated with vocabulary test performance.

**Method:** 2,181 English-speaking adults aged 18–89 completed 24 four-alternative synonym items administered as a 1-up/1-down staircase over 60 difficulty levels, with step sizes that started large (4 levels) and decreased after successive performance reversals. Two scores were compared: the QuickSIN estimate over all trials (the default) and the mean of the last four difficulty levels. Retests were available at 1 day in 100 participants, at 12 months in 307; reliability was assessed with Pearson and intraclass correlations. Twenty-one demographic correlates were screened in the 2,176 participants with questionnaire data, and a seven-term comprehensive regression model was compared with age–education–gender (AEG) adjustment, each with and without race indicators. A dichotomous Rasch model was fitted to the trial-level responses to calibrate the difficulty assignments of the stimulus set.

**Results:** Median administration time was 227 s. QuickSIN and last-four scores agreed closely (r = 0.98) and retested at 0.83 and 0.82 at 1 day and 0.81 and 0.79 at 12 months. Education correlated r = 0.43 with the vocabulary score, weekly reading hours r = 0.39, age r = 0.29, highest household income r = 0.27, and parental education r = 0.26; English as a second language correlated r = −0.19; gender did not (r = 0.003). Vocabulary scores were 0.15 levels higher per year of age across the cross-sectional sample with improvements seen in every education stratum. The comprehensive model explained 32% of the variance (10-fold cross-validated R^2^ = .31; AEG, 17% in the same participants) and attenuated but did not eliminate group differences: adjusted coefficients relative to non-Hispanic White participants were −0.27 SD for Hispanic, −0.38 for non-Hispanic Asian, −0.38 for non-Hispanic “other”, and −0.76 for non-Hispanic Black participants, all significant. Word difficulties calibrated separately in each group agreed with the non-Hispanic White calibration at r = 0.96. Rasch calibration gave ρ = 0.95 between assigned and estimated difficulty among the 316 well-sampled words, with a median displacement of 2 levels and a within-level spread of 2.9 levels; rescoring on the calibrated scale raised 12-month retest reliability to 0.84. Calibrated difficulty correlated ρ = −0.85 with Brysbaert et al. (2019) word prevalence norms.

**Conclusions:** The CCAB vocabulary test provides a reliable four-minute estimate of verbal ability that increased with age. The assigned difficulty levels were broadly correct, but the spread of difficulty within a level limited the resolution of level-based scores to approximately 2–3 levels.

Key Findings

- A 24-item adaptive vocabulary test, stepping over 60 difficulty levels (10 possible words per level) was completed by 2,181 English-speaking adults aged 18–89 in a median of 227 s.
- QuickSIN-style vocabulary scores based on all 24 trials retested at 0.83 (1 day) and 0.81 (12 months); the average difficulty level of the last 4 trials retested at 0.82 and 0.79.
- Education was the strongest demographic correlate (r = 0.43), followed by weekly reading hours (r = 0.39), age (r = 0.29), and English as a second language (r = −0.19). There was no gender difference (d = 0.007).
- Vocabulary scores increased with age across the cross-sectional sample (29.8 at 18–29 to 39.0 at 75–89, 0.15 levels per year of age, d = +1.00) within every education stratum.
- A seven-term comprehensive model (age, education, reading, computer use, English as a second language, parental education, and household income) explained 32% of the variance (10-fold cross-validated R^2^ = .31) against 17% for age–education–gender adjustment in the same participants. The comprehensive model attenuated but did not eliminate the racial/ethnic differences: with race/ethnicity indicators added, the adjusted coefficients relative to non-Hispanic White participants were −0.27 SD for Hispanic, −0.38 for non-Hispanic Asian, −0.38 for non-Hispanic “other”, and −0.76 for non-Hispanic Black participants.
- A Rasch calibration of item difficulty from trial-level responses strongly supported the assigned difficulty levels (ρ = 0.95 among the 316 words tested at least 50 times) but found that words sharing a level were spread across about 2.9 levels of difficulty, so the effective resolution of a level-based score was approximately 2–3 levels. Rescoring on the calibrated scale raised 12-month retest reliability from 0.81 to 0.84 (p < .001).
- Rasch difficulties estimated separately in NH Black, NH Asian, and Hispanic participants agreed with the NH White calibration at r = 0.96 across the 91–102 words well sampled in each pair.
- Against the independent word prevalence norms of Brysbaert et al. (2019), covering 499 of the 593 administered words, calibrated Rasch difficulty correlated ρ = −0.85 and the assigned level ρ = −0.87.

## Introduction

Vocabulary is among the most stable indices of verbal ability in the adult neuropsychological repertoire (Yuspeh et al., 1998). It resists the decline seen in speeded and fluid measures, permitting the estimation of premorbid ability and improving the classification of impairment when it is included in demographic correction (Lezak et al., 2012).

Vocabulary tests can be broadly divided into three formats: (1) fixed-form tests in which all participants receive the same set of items; (2) terminating fixed-form tests in which all participants receive the same predetermined sequence that halts based on discontinuation rules; and (3) adaptive tests in which item difficulty changes dynamically as a function of the participant’s responses. Most clinical vocabulary measures use one of the first two approaches. Vocabulary tests incorporate four major response formats: (1) Verbal definitions scored by the examiner (e.g., WAIS); (2) Pronunciation of irregular words, scored by the examiner (e.g., NART, ANART, TOPF); (3) Picture-word matching scored objectively (PPVT, NIH toolbox); and (4) Synonym selection, scored objectively.

For example, a fixed-form verbal test with stopping rules, such as the WAIS Vocabulary subtest (Wechsler et al., 2008), moves a participant through thirty items from a designated start point and stops after three consecutive failures. Several features limit the precision of this approach. First, response scoring depends partly on subjective judgment and may therefore be influenced by the concordance of the participant’s response style, dialect, and strategy with that of the examiner. Second, while items may be reliably ordered by global difficulty at a population level, ordering may not hold for the individual words at a particular difficulty level in the test because an individual’s word knowledge is influenced by education, occupation, reading experience, culture, and idiosyncratic exposures. Third, scores reflect response variability as well as underlying vocabulary knowledge: an anomalous failure on an easy item affects the score even when it provides little information about the boundary of the individual’s vocabulary. Finally, fixed-sequence testing is inefficient, administering items below the participant’s ability level, while undersampling vocabulary around the critical boundary – at and above the estimated vocabulary level.

Irregular-word reading tests such as the National Adult Reading Test (Nelson & Willison, 1982), the AmNART (Grober et al., 1991) and the Test of Premorbid Functioning [TOPF] (Wechsler, 2009) require individuals to read aloud irregular words with their pronunciation evaluated by the examiner. Irregular-word pronunciation tests measure *familiarity* with the conventional pronunciation of words rather than their semantic representations. Consequently, a native speaker may have encountered a word often enough to pronounce it correctly without knowing its precise meaning. Conversely, an individual—particularly a non-native English speaker—may understand a written word while being unfamiliar with its conventional English pronunciation. Pronunciation-based scoring may also be influenced by a participant’s accent or dialect. Moreover, ceiling effects are common, limiting sensitivity in individuals with large vocabularies (van der Linde & Bright, 2024).

Thus, performance on tests such as the NART shares about 50% of variance (r = 0.75) with WAIS vocabulary scores (Uttl, 2002).

Picture-vocabulary tests such as the Peabody Picture Vocabulary Test (Dunn & Dunn, 2007) and NIH Toolbox Picture Vocabulary Test (Gershon et al., 2014) require participants to match a picture to a word and thus provide an efficient, objectively scored measure of receptive vocabulary. The NIH Toolbox Picture Vocabulary Test is a well-calibrated example: its 602-word bank was fit with a one-parameter Rasch model on 4,703 participants (including 1,513 adults) tested through 21 overlapping forms of 40 to 60 items, with at least 200 responses per item (Gershon et al., 2014). Importantly, it is administered adaptively, stopping at 25 items or when the standard error of the ability estimate falls below 0.3, and retests with an intraclass correlation of .80 and a convergent validity of r = .80 against the PPVT-4 (Dunn & Dunn, 2007). However, picture vocabulary word selection is constrained by the imageability requirement, with high concreteness words constituting about 20% of large word corpora (Brysbaert et al., 2014). Because every target and distractor must be represented unambiguously by an image, picture-vocabulary tests necessarily limit the representation of abstract, relational, and semantically nuanced vocabulary.

Synonym matching tests require participants to select synonyms or phrases most closely matching the meaning of a word. For example, the Shipley vocabulary test is a self-administered test of 40 words where participants select the closest synonym from four alternatives (Kaya et al., 2012). More recent vocabulary tests using synonym selection have incorporated larger corpora. For example, the Vocabulary Size Test (Nation & Beglar, 2007) orders items purely by word frequency band (ten items per 1,000-word-family band over fourteen bands of the British National Corpus) with no empirical difficulty estimates and no adaptive rule. LexTALE (Lemhöfer & Broersma, 2012) uses word/non-word discriminations, selecting 60 items from a 240-item pool using classical percent-correct values and item–total correlations obtained from a pilot of eighteen participants, and then deliberately did not order them by difficulty, arranging them instead so that no more than five words or nonwords appeared consecutively. Recently, the NIH Mobile Toolbox group developed a Word Meaning test (Young et al., 2024) incorporating more than 1,000 words using norming procedures similar to those used to develop the Picture Vocabulary test. The Word Meaning test properties are compared with those of the CCAB vocabulary test in the discussion below.

The CCAB vocabulary test uses a 4-choice synonym selection test based on a difficulty ordering derived from aggregated response data from the widely used, publicly accessible online vocabulary game FreeRice.com, in which players selected word synonyms. We categorized 600 ranked words into 60 levels, edited the synonym alternatives, and reordered the list with Rasch procedures against pilot data in 60 participants.

## Method

### Participants

There were 2,181 English-speaking adults who completed the vocabulary test between May 2021 and September 2026. The CCAB Spanish-language vocabulary test uses a separate word bank and was excluded. Relatively complete questionnaire and demographic data were available for 2,176 participants: age 18 to 89 years (M = 51.9, SD = 17.4); 57.4% of the 2,142 who reported sex were women; mean education was 15.4 years (SD = 2.4). Race/ethnicity was coded as a mutually exclusive variable; of the 2,175 reporting race there were 367 Hispanic/Latino participants of any race (the coding used in the CCAB C-model), 709 non-Hispanic (NH) Whites, 475 NH Blacks, 388 NH Asians, and 236 NH “other”, including mixed race. Age-band sizes were 291 at 18–29, 505 at 30–44, 526 at 45–59, 659 at 60–74, and 195 at 75–89. Three hundred participants (13.8%) reported that English was not spoken in their home while growing up. Testing was telemedical: participants were tested in their homes by an experienced proctor who monitored performance through a built-in web interface. All participants gave written informed consent under WIRB protocol 20201196. The vocabulary test is one subtest of the California Cognitive Assessment Battery (Woods et al., 2024); a companion report describes the battery’s scoring models which include the vocabulary score as a covariate in the comprehensive C-model correction of the other tests (Woods et al., 2026). Table 1 summarizes the sample.

**Table 1.**
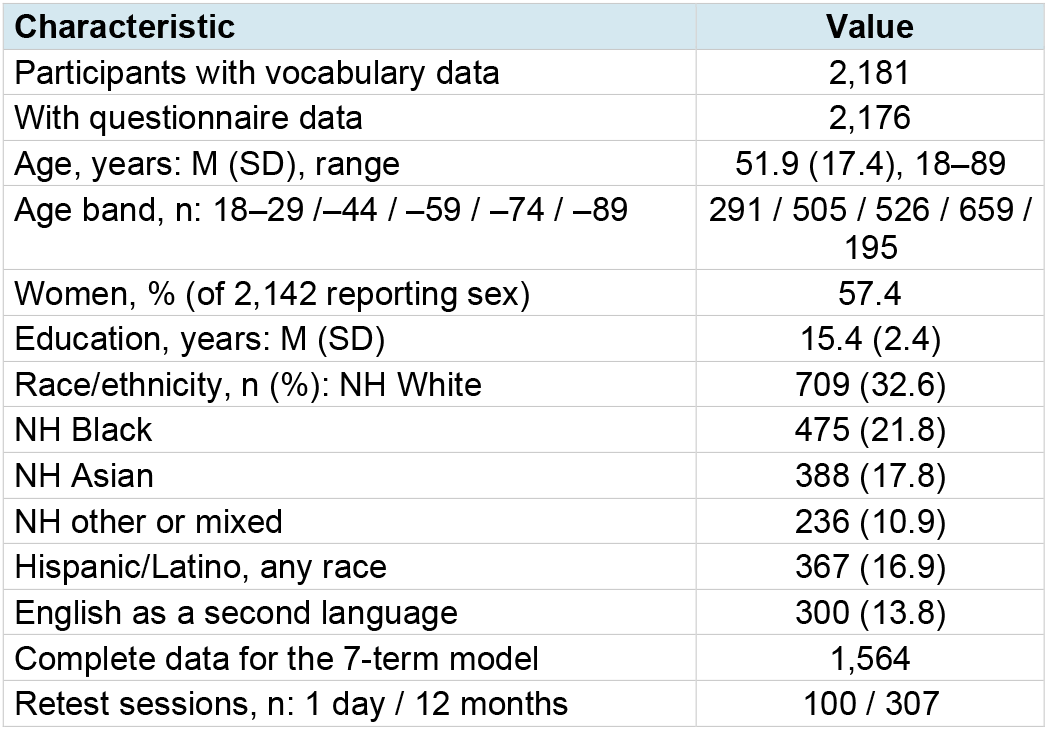
Participants. Note. Race/ethnicity percentages are of the 2,175 participants reporting race; Hispanic/Latino participants of any race are classified as Hispanic and the remainder by race (NH = non-Hispanic). Complete questionnaire data (age, sex, education, race, ethnicity, and language background) were available for 2,176 of the 2,181 participants.

### The word bank

The 600-word bank, with 10 words at each of 60 difficulty levels, was derived from aggregated response data from the online vocabulary game FreeRice.com, in which players answered four-alternative synonym questions.

The list was pruned, edited, and reordered with Rasch procedures against pilot CCAB data and edited prior to use with the present sample. Each item presents a target word with four response alternatives, one of which is a synonymous word or short phrase.

### The adaptive vocabulary test

Participants responded by screen touch with 24 words presented to every participant. There was no time limit, and administration was self-paced. The task, including instructions, required a median of 227 s (IQR 200–258 s); the interval from the first stimulus to the last response had a median of 173 s (IQR 146–203 s).

Difficulty was adapted with a 1-up/1-down staircase: a correct response moved the participant to a harder level and an error to an easier one. Participants began at level 16 with step sizes of 4, 4, 3, 3, 2, 2, and 1, with the step size decreasing after successive performance reversals (a change from correct to incorrect or vice versa). These large initial step sizes generally brought participants near their final difficulty level within 10–18 trials (Figure 1). Duplicate word selection at a repeated level (e.g., level 34 in Figure 1) was avoided by sampling words at that level without replacement.

**Figure 1.**
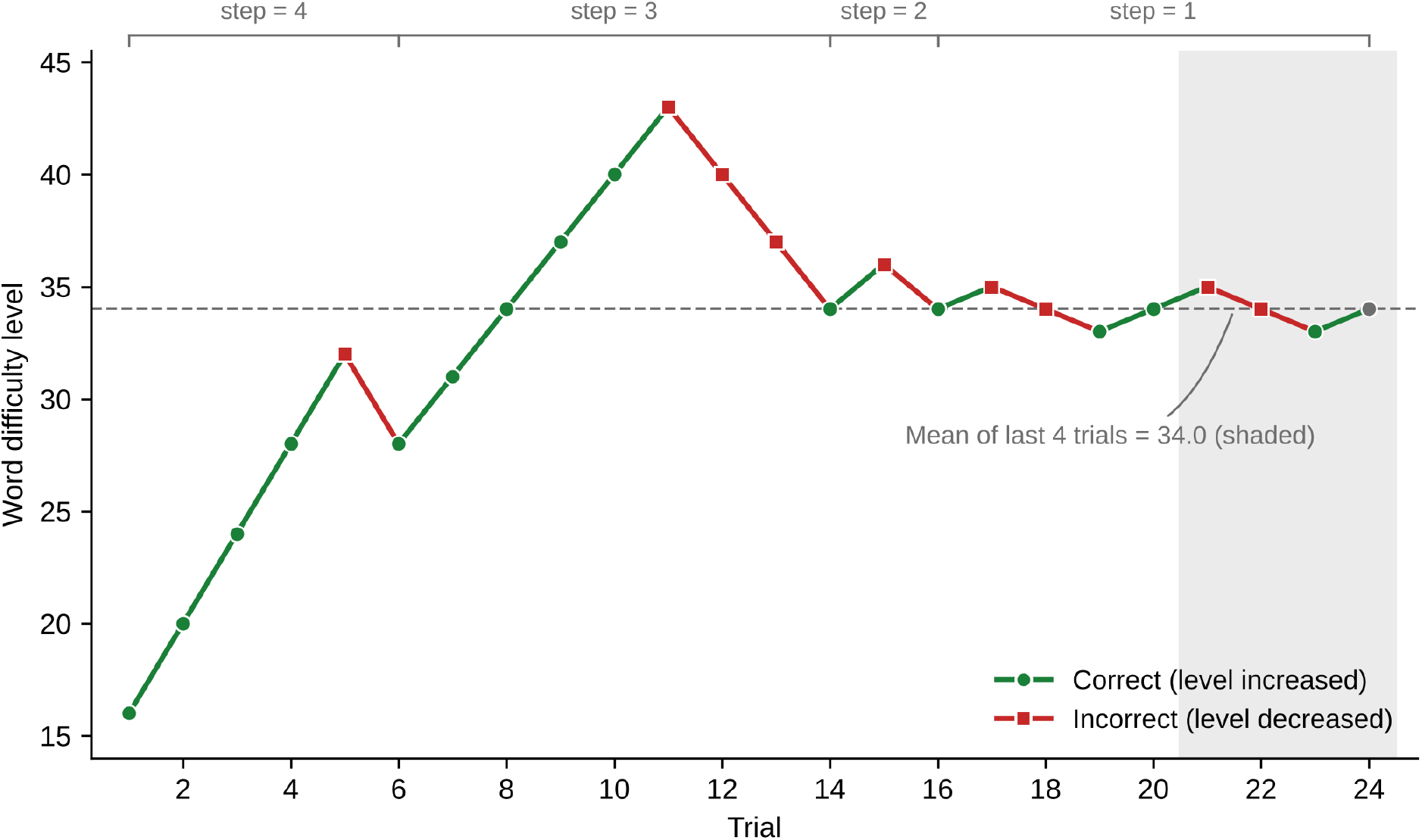
Adaptive staircase trajectory for a representative participant. Testing began at level 16. Correct responses raised the difficulty level (green) and incorrect responses lowered it (red); the step size declined from 4 to 1 level after successive performance reversals (annotated above), initially overshooting the final vocabulary level (step = 3). This participant was within one level of the final value by trial 14. The dashed line marks the mean of the final four levels (shaded). Across the eight trials administered at the smallest step size, the participant answered 50% correctly.

### Scoring

Two vocabulary scores were computed. The default was the QuickSIN estimate, the Spearman–Kärber arithmetic used to score the QuickSIN speech-in-noise test (Killion et al., 2004). The proportion correct was computed at each difficulty level visited, so that all 24 trials contributed, and the score was the Spearman– Kärber estimate of the level at which the proportion correct crosses 0.5. With the levels visited ordered L_1_ < … < L_k_ and p_i_ the proportion correct at L_i_, the proportion was taken as p_0_ = 1 at L_0_ < L_1_ − 1 and as p_k+1_ = 0 at L_k+1_ > L_k_ + 1, and the score is Σ_i_ (p_i_ − p_i+1_)(L_i_ + L_i+1_)/2 for i = 0 … k, the weighted estimate of the 50%-correct level. An alternative score, the mean of the last four difficulty levels, was closely related (r = 0.98). Observed ranges were 3.75 to 57.75 for the QuickSIN estimate and 1.00 to 59.75 for the mean of the last four. Throughout this report, “difficulty level” refers to a word’s position in the bank and “vocabulary score” to a participant’s estimate on the same 1–60 scale. Both scores were also compared with the model-based ability estimate from the Rasch calibration described below.

Accuracy, the proportion of the 24 items answered correctly, reflected how quickly the adaptive staircase settled rather than where it settled. Response times were summarized as the mean of log response time over responded trials, for correct trials and for all trials, and as the coefficient of variation of the raw times.

### Demographic screen and models

Twenty one candidate demographic correlates were screened: age, education in years, gender, Latino ethnicity, daily medication count, a socioeconomic composite, computer use, weekly hours of reading, parental education for each parent and their mean, highest and most recent household income, subjective social standing (a 0–1 self-rating), number of rooms in the household, marital status (married vs. not), whether English was spoken in the home while growing up (coded as English as a second language, ESL, when it was not), family history of dementia, and the GAD-7, GDS, and Cognitive Failures Questionnaire scales. Self-reported race and Hispanic ethnicity were combined into one mutually exclusive race/ethnicity variable — Hispanic/Latino of any race, then non-Hispanic (NH) White, Black, Asian, and other or mixed race — the coding used in the CCAB C-model (Woods et al., 2026), and analyzed as group means and as model indicators rather than as a screened correlate. Of the 367 Hispanic participants, 256 recorded their race as other. Education in years was coded from the categorical education item by the standard CCAB mapping; the four education categories used in the C-model (high school or less, some college or associate’s degree, bachelor’s degree, graduate degree) were examined as an alternative specification. Age-squared was orthogonalized against age.

Two regression models of the vocabulary score were compared. The first was the common age–education– gender (AEG) adjustment in clinical use. The second, the comprehensive model, was selected from the 21 demographic and questionnaire candidates by iteratively removing the predictor with the smallest standardized coefficient until all remaining |β| ≥ 0.10, on the 1,420 participants with all candidate predictors. This retained eight terms. Most recent household income (β = −0.12, zero-order r = +0.18) was then dropped because its negative weight arose only in the presence of highest household income, leaving seven terms: age, education, reading hours, computer use, ESL, parental education, and highest household income. After selection, the seven-term model was refitted in the 1,564 participants with complete data on the seven predictors. Each model was fitted with and without race/ethnicity indicators (NH Black, Hispanic, NH Asian, and NH other, against NH White). Residual scores were compared across the five groups, and the proportion of each group falling in the bottom 7% of residuals in the entire population, the CCAB low-score flag, was tabulated. The comprehensive model was checked by 10-fold cross-validation and by bootstrap estimates of coefficient stability.

### Rasch calibration of the word bank

A dichotomous Rasch model was fitted to the trial-level responses of all 2,181 participants to estimate each word’s difficulty independently of its assigned level. The model is P(correct) = 1 / (1 + exp(−(θ − b))), with θ the examinee’s ability and b the word’s difficulty on a common logit scale. It was fitted by marginal maximum likelihood with an EM algorithm over a 71-node quadrature, using each participant’s first session (52,224 responses). The current operational bank contains 600 words. Eleven retired items that appeared transiently during a 2022–2023 bank revision (121 responses in all) were excluded from calibration and scoring; of the 600 current-bank words, 593 were administered at least once and seven, at levels 2, 3, and 59, were never sampled. A weak empirical-Bayes prior, b ∼ N(α + β × assigned level, τ^2^) with the hyperparameters estimated from words answered at least 50 times, kept sparsely sampled words finite and pulled them toward the difficulty typical of their assigned level. Every session was then scored on the frozen word difficulties with Warm’s weighted likelihood estimate of ability (Warm, 1989). The within-level spread of difficulty was summarized as the SD of the calibrated difficulties of well-sampled words around their assigned level’s mean, corrected for sampling variance, and expressed in levels by dividing by the slope of difficulty of assigned levels.

To test whether the word difficulties were the same across race/ethnicity groups, the model was refitted separately to the NH White, NH Black, NH Asian, Hispanic, and NH other participants (the mutually exclusive coding above). Each group’s scale was linked to the NH White scale by equating the mean difficulty of the words answered at least 30 times in both groups, and differential item functioning was computed for each of those words as the group minus NH White difficulty on the linked scale, with a false-discovery-rate correction over words within each group. The SD of the differential item functioning expected from estimation error alone was the root mean square of the standard errors of the paired difficulty estimates. Because the items are four-alternative, the calibration was repeated as a sensitivity analysis with a fixed lower asymptote of 0.25, P = 0.25 + 0.75 × [1 + exp(−(θ − b))]−^1^, and the two solutions were compared on item ordering, person scores, and retest reliability.

As an external criterion, the calibrated difficulties and the assigned levels were compared with the word prevalence norms of Brysbaert et al. (2019), the proportion of 221,268 respondents in a lexical-decision task who reported knowing each of 61,855 English lemmas, expressed on a probit scale.

### Reliability

Retests occurred at 1 day (n = 100) and 12 months (n = 307) for different participants. Retest reliability was computed as the Pearson correlation and as the intraclass correlation for absolute agreement from a two-way random-effects model, ICC(A,1), with 95% confidence intervals (McGraw & Wong, 1996). Dependent correlations measured in the same participants were compared with Steiger’s test.

## Results

Figure 2 shows the score distribution, accuracy, and a comparison of the two scoring measures. The distribution of QuickSIN scores was close to symmetric (M = 34.37, SD = 9.20, median 35.13, skew −0.38) and spanned most of the 60-level range, from 3.75 to 57.75. Floor and ceiling effects were negligible: one participant reached a mean of the last four of 59.75 and one a mean of 1.00. The two scores (Figure 2, right panel) behaved almost identically in every analysis, as expected from the high correlation (r = 0.98; SD of the difference = 1.94), with the QuickSIN estimate lower by 0.36 levels due to occasional misses early in the sequence.

**Figure 2.**
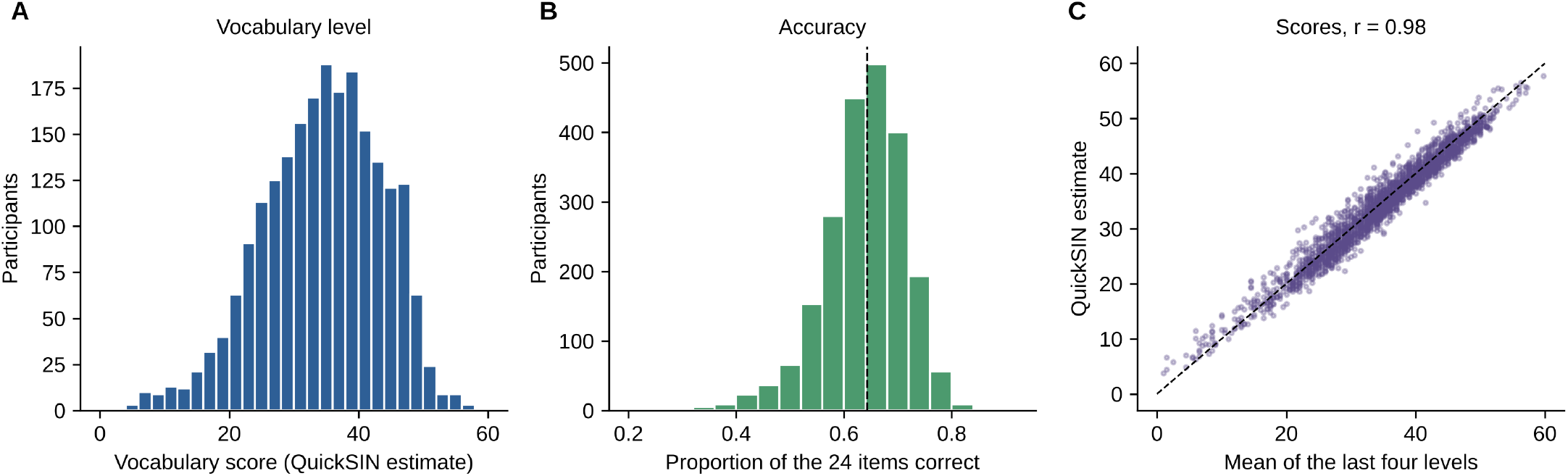
Vocabulary level, accuracy, and scores. (A) Distribution of the vocabulary level (QuickSIN estimate) across the 2,181 participants, spanning most of the 60-level range with few participants near either end. (B) Distribution of accuracy, the proportion of the 24 items answered correctly; the dashed line marks the sample mean of 0.64. (C) The QuickSIN estimate versus the mean of the last four difficulty levels (r = 0.98); the dashed line is the identity.

The QuickSIN estimate retested at 0.83 at 1 day and 0.81 at 12 months against 0.82 and 0.79 for last-four mean. The insignificant difference ran in the direction theory predicts, since a score that uses all twenty-four trials should be less exposed to noise in the last few trials than one that uses four. Both scores correlated 0.99 with the Rasch ability estimate derived from the same responses (QuickSIN 0.992, last four 0.985), so both scores closely tracked the model-based ability estimate.

Table 2 summarizes the results and the correlations with age, education, and gender predictors as well as test-retest reliability at 1 day and 12 months. Mean accuracy was 0.644 (SD 0.080, IQR 0.583–0.708), corresponding to 15.4 of 24 items correct. Accuracy fell across trials as the staircase approached each participant’s threshold, from 0.92 on trial 1 to 0.55 over the final four trials, consistent with the 50%-correct convergence point of a symmetric up–down staircase. Accuracy correlated r = 0.66 with the vocabulary score and retested at 0.24 and 0.35.

**Table 2.** Measure screen. The two vocabulary scores, the Rasch ability estimate, and four secondary measures, with distribution, age, education and sex effects, model fit, and retest reliability. Note. Distributions are for all 2,181 participants; age, education, sex, and model columns use the 2,176 with demographic data. Response times are the means of log response time over responded trials; M is the geometric mean and SD is M × (exp(SD of the logs) − 1). d age contrasts the 75–89 and 18–29 bands. R^2^ comp is the seven-term comprehensive model and R^2^ AEG the age–education–gender model, both on the 1,564 participants with complete data for the seven terms. Retest n = 100 at 1 day and 307 at 12 months; r is the Pearson correlation, and the intraclass correlation for absolute agreement equals r to two decimals for the vocabulary scores and accuracy (see Results).

| Measure | n | M | SD | r age | d age | r edu | d sex | R<sup>2</sup> comp | R<sup>2</sup> AEG | r 1d | r 12mo |
| --- | --- | --- | --- | --- | --- | --- | --- | --- | --- | --- | --- |
| Vocabulary QuickSIN estimate | 2,181 | 34.37 | 9.20 | 0.29 | +1.00 | 0.43 | +0.007 | 0.32 | 0.17 | 0.83 | 0.81 |
| Vocabulary score (mean last 4) | 2,181 | 34.73 | 9.36 | 0.28 | +1.01 | 0.42 | -0.007 | 0.30 | 0.17 | 0.82 | 0.79 |
| Rasch ability (logits) | 2,181 | -0.246 | 1.77 | 0.29 | +1.03 | 0.43 | +0.007 | 0.32 | 0.17 | 0.84 | 0.84 |
| Accuracy | 2,181 | 0.644 | 0.080 | 0.18 | +0.63 | 0.30 | -0.048 | 0.13 | 0.07 | 0.24 | 0.35 |
| Items correct (of 24) | 2,181 | 15.45 | 1.93 | 0.18 | +0.63 | 0.30 | -0.048 | 0.13 | 0.07 | 0.24 | 0.35 |
| Response time, mean (ms) | 2,181 | 5457 | 2026 | 0.13 | +0.49 | -0.13 | -0.106 | 0.05 | 0.04 | 0.71 | 0.68 |
| Response time, CoV | 2,181 | 0.453 | 0.116 | -0.16 | -0.48 | 0.12 | +0.180 | 0.06 | 0.04 | 0.46 | 0.39 |

### Demographic correlates

Figure 3 shows the effects of demographic predictors on vocabulary. Education was the strongest predictor (r = 0.43, n = 2,175), followed by weekly hours of reading (r = 0.39), age (r = 0.29), highest household income (r = 0.27), parental education (r = 0.26 for the mean of both parents), and computer use (r = 0.21). English as a second language (300 participants, 13.8%) correlated r = −0.19 (n = 2,176): participants who did not speak English at home growing up averaged 29.9 against 35.1 for those who did (d = −0.57). Most recent household income was a weaker correlate than highest household income (0.18 against 0.27), consistent with vocabulary tracking lifetime rather than current circumstances, as many older participants were retired.

**Figure 3.**
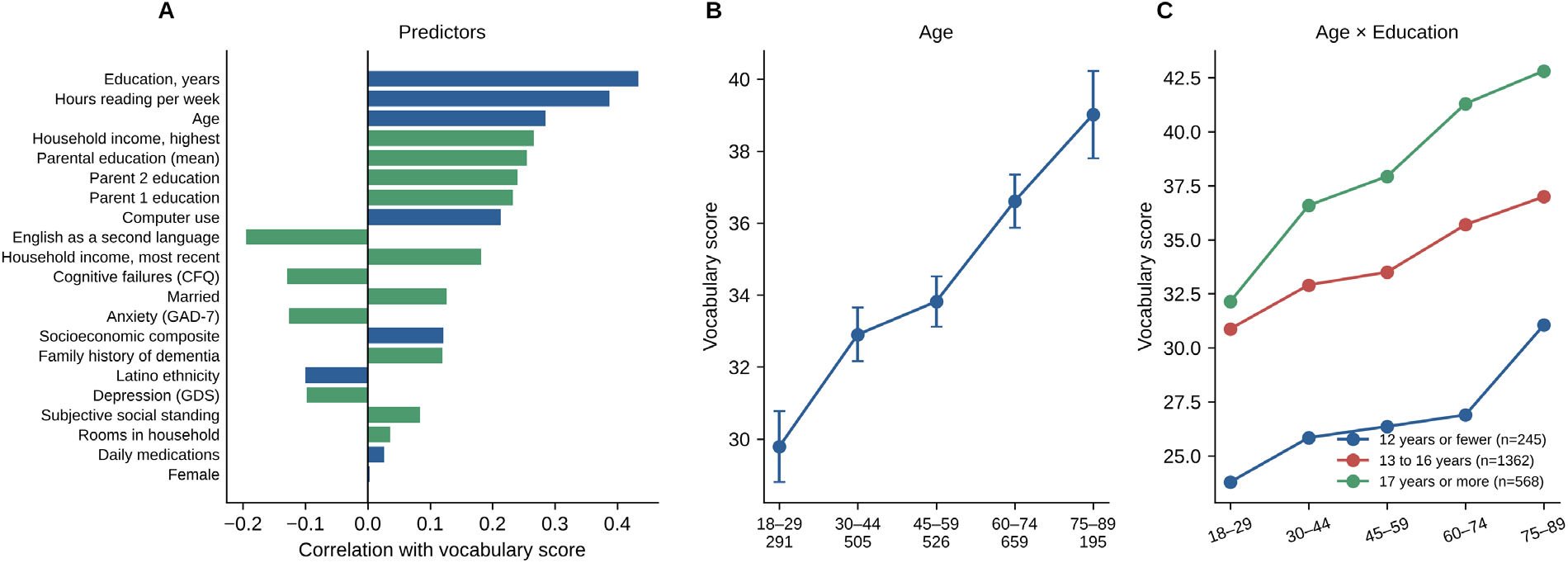
Demographic predictors of vocabulary. (A) Predictors: zero-order correlations of the vocabulary score with the twenty-one screened variables, ordered by magnitude; the eight variables carried in the test export are blue and the thirteen questionnaire variables green. (B) Age: vocabulary score by age band, with 95% confidence intervals and band sizes below each point. (C) Age × Education: the same age curve within three education strata.

As seen in Figure 3, vocabulary rose monotonically across the five age bands (r = 0.29, 0.15 levels per year; d = +1.00 between the oldest and youngest bands). The quadratic term was not significant (t = 1.15, p = .25); no departure from linearity was detected across the span sampled, and no downturn in the oldest band. Within each of three education strata the age gradient persisted: r = 0.18 among the 245 participants with 12 years or fewer, r = 0.21 among the 1,362 participants with 13 to 16 years, and r = 0.35 among the 568 participants with 17 years or more. The youngest-to-oldest gain within strata was 7.3, 6.1, and 10.7 levels respectively, with the largest gain among those with the most education.

Table 3 and the left panel of Figure 3 show the influence of the 21 screened demographic correlates, ordered by the magnitude of their correlation with the vocabulary score. Education, reading and age were correlated with one another, so each was tested with the other two removed (Table 4). All three survived. Education retained a partial r of 0.34 with reading and age controlled (p < .001, n = 2,147); reading retained 0.28 with education and age controlled; and age retained 0.20 with education and reading controlled. English as a second language retained a partial r of −0.26 with education and age controlled (p < .001, n = 2,175). Parental education retained a partial r of 0.24 with the participant’s own education and age removed (p < .001, n = 1,578), so the family-of-origin contribution was not merely a proxy for attainment. Cumulatively, age alone accounted for 8% of the variance in the vocabulary score, age, education and gender (AEG) for 24% in the full sample, AEG plus reading for 29%, and AEG, reading and ESL for 34% (n = 2,113).

**Table 3.** Zero-order correlations of the screened demographic and questionnaire variables with the vocabulary score. Note. The parental-education, income, and social-standing items were completed by about three-quarters of the sample, while the remaining questionnaire items are essentially complete. English as a second language = English not spoken in the home while growing up (300 of 2,176). Married and family history of dementia are age-confounded and do not survive adjustment for age and education (see Results).

| Variable | $r$ | $p$ | $n$ |
| --- | --- | --- | --- |
| Education, years | 0.43 | < .001 | 2,175 |
| Hours reading per week | 0.39 | < .001 | 2,147 |
| Age | 0.29 | < .001 | 2,176 |
| Household income, highest | 0.27 | < .001 | 1,628 |
| Parental education (mean) | 0.26 | < .001 | 1,578 |
| Parent 2 education | 0.24 | < .001 | 1,491 |
| Parent 1 education | 0.23 | < .001 | 1,557 |
| Computer use | 0.21 | < .001 | 2,156 |
| English as a second language | -0.19 | < .001 | 2,176 |
| Household income, most recent | 0.18 | < .001 | 1,628 |
| Cognitive failures (CFQ) | -0.13 | < .001 | 2,174 |
| Married | 0.13 | < .001 | 2,121 |
| Anxiety (GAD-7) | -0.13 | < .001 | 2,171 |
| Socioeconomic composite | 0.12 | < .001 | 2,173 |
| Family history of dementia | 0.12 | < .001 | 1,981 |
| Latino ethnicity | -0.10 | < .001 | 2,168 |
| Depression (GDS) | -0.10 | < .001 | 2,173 |
| Subjective social standing | 0.08 | < .001 | 1,540 |
| Rooms in household | 0.04 | .090 | 2,174 |
| Daily medications | 0.03 | .224 | 2,165 |
| Female | 0.00 | .873 | 2,142 |

**Table 4.** Partial correlations with the vocabulary score.

| Predictor, with the stated variables removed | r | p | n |
| --- | --- | --- | --- |
| Education, controlling reading and age | 0.34 | < .001 | 2,147 |
| Reading, controlling education and age | 0.28 | < .001 | 2,147 |
| English as a second language, controlling education and age | -0.26 | < .001 | 2,175 |
| Parental education, controlling own education and age | 0.24 | < .001 | 1,578 |
| Age, controlling education and reading | 0.20 | < .001 | 2,147 |

Three affective and self-report scales correlated negatively and weakly: cognitive failures (CFQ, r = −0.13), anxiety (GAD-7, r = −0.13), and depression (GDS, r = −0.10). Marital status (married vs. not, r = 0.13) and a reported family history of dementia (r = 0.12) correlated positively, but both groups were older than their comparison groups (married 58.5 vs. unmarried 49.1 years; family history 58.2 vs. 50.5 years), and neither correlation survived adjustment for age and education (married, partial r = 0.00; family history, partial r = 0.05). Men and women performed similarly: the zero-order correlation with female sex was r = 0.003 (p = .87, n = 2,142), d = 0.007 (95% CI −0.08 to +0.09). Women responded slightly faster (t = −2.42, p = .015) and were more variable in their response times (t = 4.13, p < .001), but these are properties of a self-paced lexical decision rather than of vocabulary.

Education’s effect was concave: mean scores were 26.2 for high school or less (n = 245), 32.1 for some college (n = 741), 35.8 for a bachelor’s degree (n = 621), and 39.4 for a graduate degree (n = 568), steps of 0.64, 0.41, and 0.39 SD. Replacing education in years with the four C-model categories did not significantly raise R^2^ and left the other coefficients unchanged (high school or less −0.81, some college −0.33, and graduate degree +0.16 SD relative to a bachelor’s degree), so the linear term was retained here.

Racial and ethnic differences were among the largest seen with any CCAB test: NH White 39.7 (n = 709), NH other or mixed 33.5 (n = 236), NH Asian 33.0 (n = 388), Hispanic 32.3 (n = 367), and NH Black 29.6 (n = 475), giving effect sizes against the NH White mean of d = −0.67, −0.73, −0.80, and −1.10 (SD of the full sample). The Black–White difference was present in every education stratum (12 years or fewer: 22.9 vs. 32.1; 13–16 years: 30.0 vs. 38.6; 17 or more years: 35.5 vs. 43.0) and in every age band (24.4 vs. 35.5 at 18–29 through 32.5 vs. 42.9 at 75–89). It was not a language-background effect: NH Black participants were the least likely to report ESL (1.9%, against 4.7% of NH White, 8.1% of NH other, 24.3% of Hispanic, and 38.7% of NH Asian participants), and among native English speakers only, the difference was d = −1.10. It was of the same size on the Rasch ability scale, so it did not depend on the staircase scoring.

Table 5 gives the comprehensive model with and without race/ethnicity indicators, for the 1,564 participants with complete data for its seven terms. Without race/ethnicity, the seven terms explained 32% of the variance against 17% for AEG in the same participants (10-fold cross-validation of the final seven-term model, R^2^ = 0.31 and 0.17). ESL carried the largest weight (β = −0.54, a group difference in SD units), followed by education (0.28), reading (0.19), age (0.15), parental education (0.15), highest household income (0.10), and computer use (0.10); all bootstrap confidence intervals excluded zero. Adding the race/ethnicity indicators raised R^2^ to 38% and left the seven coefficients essentially unchanged, so the covariates were not proxies for race/ethnicity; the terms were NH Black β = −0.76 (t = −12.9), NH other −0.38, NH Asian −0.38, and Hispanic −0.27 (t = −4.2), each against NH White.

**Table 5.** The comprehensive demographic model without (left) and with (right) race/ethnicity indicators. Coefficients (β) in SD units of the vocabulary score, with t statistics. Note. Both models were fitted on the 1,564 participants with complete data for the seven terms. Continuous predictors were standardized; ESL and the four race/ethnicity terms are 0/1 indicators, so their coefficients are group differences in SD units of the vocabulary score. Columns 2–3: seven-term model; columns 4–5: the same model with race/ethnicity indicators against NH White. In raw units, Vocabulary score = 4.36 + 0.082·Age + 1.102·Education + 0.414·Reading + 0.422·Computer use − 4.82·ESL + 1.230·Parental education + 0.545·Income; with race/ethnicity, Vocabulary score = 9.20 + 0.095·Age + 0.992·Education + 0.392·Reading + 0.483·Computer use − 5.13·ESL + 1.144·Parental education + 0.381·Income − 6.80·NH Black − 2.40·Hispanic − 3.42·NH Asian − 3.40·NH other. Age in years; education in years; reading in hours per week; computer use on a 0–7 scale; parental education as the mean of two parents on a 0–4 scale; income as the highest household income category, 0–6.

| Predictor | $\beta$ | t | $\beta$ | t |
| --- | --- | --- | --- | --- |
| Age | +0.15 | 6.7 | +0.17 | 8.0 |
| Education, years | +0.28 | 11.8 | +0.25 | 11.1 |
| Hours reading per week | +0.19 | 8.6 | +0.18 | 8.5 |
| Computer use | +0.10 | 4.3 | +0.11 | 5.2 |
| English as a second language | -0.54 | -8.8 | -0.57 | -9.3 |
| Parental education (mean) | +0.15 | 6.7 | +0.14 | 6.4 |
| Household income, highest | +0.10 | 4.4 | +0.07 | 3.2 |
| NH Black (vs. NH White) | — | — | -0.76 | -12.9 |
| Hispanic (vs. NH White) | — | — | -0.27 | -4.2 |
| NH Asian (vs. NH White) | — | — | -0.38 | -6.0 |
| NH other (vs. NH White) | — | — | -0.38 | -5.5 |
| $R^2$ | 0.32 | | 0.38 | |
| n | 1,564 |  | 1,564 |  |

The clinical consequence is reflected in the low-score flag: taking the bottom 7% of residuals as the criterion, 14.4% of NH Black and 1.8% of NH White participants were flagged under the comprehensive model (8.0:1) and 10.8% and 2.1% under AEG (5.1:1). With race/ethnicity indicators in the model the group means are equalized by construction, but 8.9% of NH Black against 3.9% of NH White participants still fell in the bottom 7% (2.3:1), because the Black residual distribution had a broader lower tail.

### Reliability

One-day retest reliability was 0.83 for the QuickSIN score and 0.82 for the mean of the last four (n = 100); at twelve months the corresponding values were 0.81 and 0.79 (n = 307). Because mean change between sessions was negligible intraclass correlations were identical to the Pearson values to two decimals: ICC(A,1) = 0.83 (95% CI 0.75–0.88) and 0.81 (0.76–0.84) for the QuickSIN score at one day and twelve months. The twelve-month ICC is numerically similar to the .80 ICC reported for the NIH Toolbox picture vocabulary test over a 7-to-21-day interval.

Practice effects were negligible, due to the large size of the item bank: with 600 words and 24 items, repeat exposure to the same word was uncommon. The vocabulary score changed by +0.19 levels at one day (d = +0.03) and +0.41 levels at twelve months (d = +0.05); neither differed from zero. Mean response time shortened by about 9% at one day and 2% at twelve months, presumably reflecting increased familiarity with the interface.

### Rasch calibration of the word bank

Because adaptive routing determines which examinees see which words, the raw percentage correct for any item is a poor measure of its difficulty. Difficulty levels 16, 20, 24, 28, and 32 were the ascending steps most examinees passed through, and their words scored 73 to 87% correct, against 54 to 70% at levels between those (i.e., that were selected following higher-level failures); a level-16 word was administered to 2,142 participants and a level-17 word to only 172. In Rasch terms the two levels were only 0.37 logits apart, about two levels’ worth of difficulty, so the accuracy gap between them (87.1% vs 59.8%) reflected the performance of different participants, not inherent item difficulty.

The assigned difficulty levels were broadly right and locally wrong (Figure 4). Assigned level and estimated difficulty correlated ρ = 0.95 among the 316 words administered at least 50 times, and ρ = 0.99 across all 600 current-bank words, the latter measure inflated by forcing the 195 words answered fewer than 20 times to their assigned levels. Ranking all words by calibrated difficulty and reassigning them ten per level, the median absolute displacement across all well-sampled words, including those that did not move, was 2 levels. Overall 40% of well-sampled words moved three or more levels, 16% moved five or more, and 4 moved ten or more. Person separation reliability was 0.93, with a mean standard error of ability of 0.48 logits. Adding the 25% guessing floor to the model changed the scale but not the ordering: it stretched the bank range from 11.2 to 13.5 logits, but the two sets of difficulties correlated r = 0.999 across the 316 well-sampled words, 99.7% of words fell within one level of the same calibrated position, person estimates correlated r = 0.997, and twelve-month retest reliability was 0.84 under both models. Because the guessing-adjusted model produced essentially identical item ordering, person ranking, and retest reliability, the simpler Rasch model was retained for the primary analyses.

**Figure 4.**
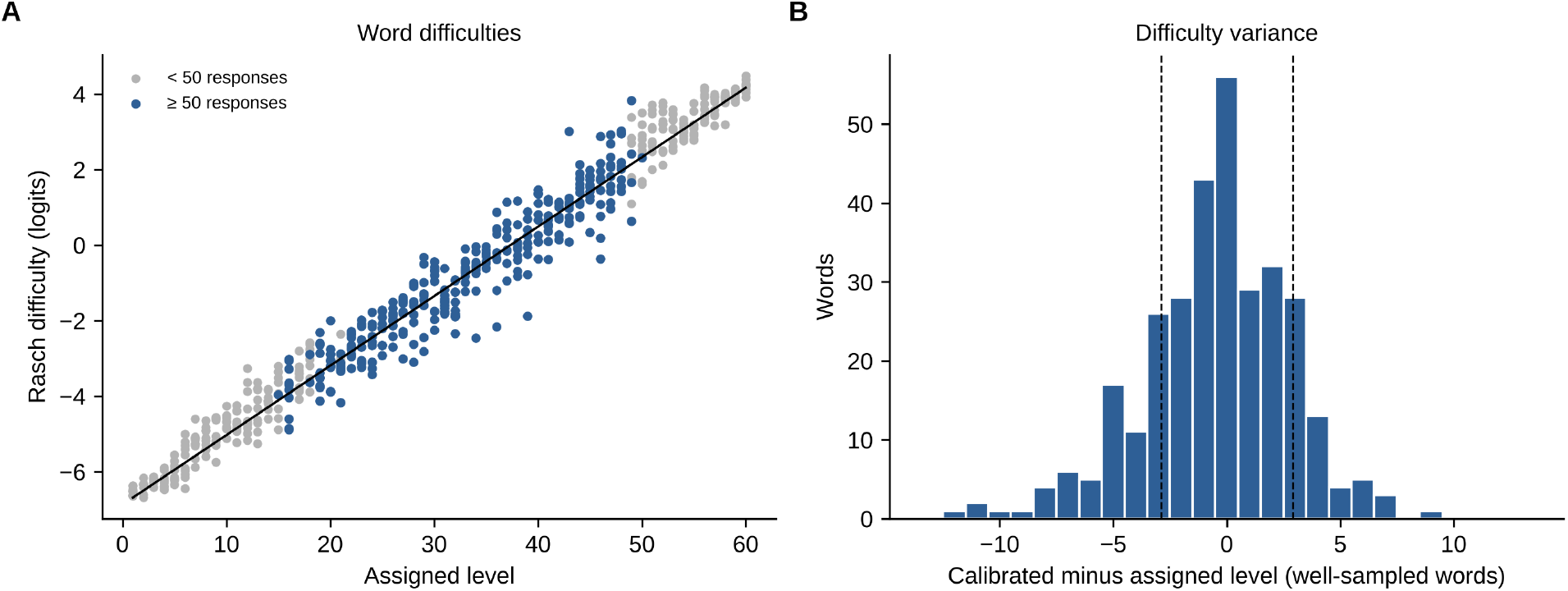
Rasch calibration of the word bank. (A) Word difficulties: estimated difficulty against assigned level for the 593 administered words, with the line fitted to the well-sampled words (0.18 logits per level); words answered fewer than 50 times are gray. (B) Difficulty variance: displacement of the well-sampled words from their assigned level, in levels; dashed lines mark ±1 within-level SD (2.9 levels).

The slope of calibrated difficulty on assigned level was 0.18 logits per level among well-sampled words. But words sharing an assigned level were spread 0.53 logits around their level’s mean. Thus, within a single level the words ranged over about 2.9 levels’ worth of difficulty. This has practical consequences: two words drawn from adjacent levels therefore differed by a mean of 0.18 logits of calibrated difficulty (SD 0.75), meaning that a one-level step actually raised the difficulty level only 60% of the time (a two-level step raised difficulty 69% of the time, and a four-level step 84%). Thus, the staircase’s smallest steps were close to a coin flip in their difficulty adjustment.

Rescoring every session on the calibrated scale left the ranking of examinees almost unchanged (r = 0.99 with both level-based scores) but improved retest reliability: at twelve months the ability estimate retested at 0.84 (ICC 0.84, 95% CI 0.80–0.87) against 0.81 for the QuickSIN score (Steiger z = 4.2, p < .001).

Fit was good: mean infit 0.90 (SD 0.10) and mean outfit 0.87 (SD 0.21) among words answered at least 30 times, with 12 words flagged as overfitting (infit and outfit both < 0.7) and 4 as underfitting (infit > 1.3 or outfit > 1.5). The largest displacements were words answered correctly more often than their assigned level implied. For example, *meme*, assigned level 39 on a list assembled before the word entered general use was answered correctly by 97% of those who saw it, moving it down twelve levels.

### Cross-group item calibration and differential item functioning

Table 6 gives the separate calibrations of each group against the 709 NH White participants. Because adaptive routing concentrates each group’s responses at different levels, only 91–102 words (levels 16–40) were answered at least 30 times in both a group and the NH White participants. Over those words the NH Black, NH Asian, and Hispanic difficulties correlated r = 0.96, 0.96, and 0.96 with the NH White difficulties, as closely as the pooled calibration agreed with the assigned levels. Before linking, each group’s difficulties were displaced throughout by its mean ability difference (1.7, 0.2, and 1.2 logits); after linking, the SD of the differential item functioning (0.44, 0.47, and 0.42 logits) matched the 0.41–0.42 expected from estimation error alone. The number of words differing at p < .05 was at chance for Black participants (5 against 5.1 expected) and modestly above it for Asian (10 against 4.6) and Hispanic (8 against 4.5) participants. Only one word in the 285 comparisons (equine, 1.3 logits harder for Black participants) survived correction for multiple comparisons. The NH other group (236 participants, 41 comparable words) was too small and heterogeneous for a stable separate calibration and is not shown. These findings are consistent with group differences reflecting displacement along a common scale, with little evidence of systematic differential item functioning in the adequately sampled portion of the bank.

**Table 6.**
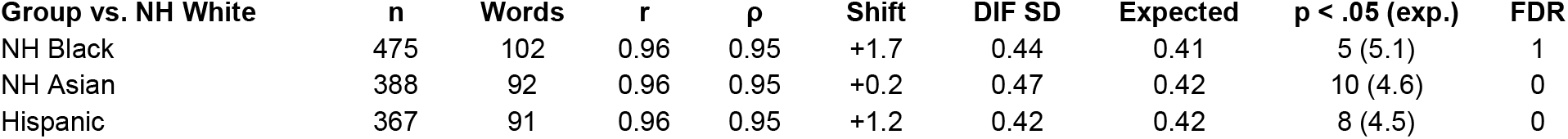
Word difficulties calibrated separately in each race/ethnicity group and linked to the NH White calibration (n = 709). Note. n = participants in the group; Words = words answered at least 30 times in both the group and NH White participants (all at levels 16–40); r and ρ = Pearson and Spearman correlations of the group’s difficulties with the NH White difficulties over those words; Shift = mean displacement of the group’s difficulties before linking, in logits (the group’s mean ability difference on the item scale); DIF SD = SD of the group-minus-White difficulty after linking, against the SD expected from estimation error alone; p < .05 = words with |z| > 1.96 (expected count under no DIF); FDR = words significant after false-discovery-rate correction (equine, for NH Black). The NH other group (n = 236, 41 comparable words) was too small for a stable separate calibration and is omitted.

### Word difficulty and word prevalence

Of the 593 administered words, 499 appear in the prevalence norms of Brysbaert et al. (2019); the 94 that did not are rare words concentrated at the hard end of the bank. Across the matched words, calibrated difficulty correlated ρ = −0.85 with prevalence and the assigned level ρ = −0.87; among the 285 well-sampled words, where the range is narrower, the correlations were −0.65 and −0.71. The proportion of respondents responding correctly fell from 1.00 at level 1 to 0.97 at level 16, 0.72 at level 31, 0.58 at level 41, and 0.31 at level 56. Prevalence added nothing to the prediction of calibrated difficulty once the assigned level was known (standardized β = 0.01 for prevalence against 0.95 for level), and within assigned levels it was unrelated to the calibrated scatter (r = 0.01). The within-level spread of difficulty therefore likely reflected properties of the synonym-choice items as well as word knowledge.

## Discussion

A 24-item adaptive test requiring four minutes produced a vocabulary score that retested at 0.83 over one day and 0.81 over one year, spanned the ability range without floor or ceiling effects, showed no practice effect, and was higher at older ages. The score was most strongly associated with education, then reading habits, age, and the indices of family and household circumstance, and non-English upbringing. A comprehensive demographic model substantially attenuated but did not eliminate the differences between NH White participants and the Hispanic, Asian, other, and Black groups; all four adjusted coefficients remained significant, the Black–White difference the largest. Rasch calibration and independent word prevalence norms strongly supported the assigned difficulty levels while showing that the resolution of a level-based score is approximately 2–3 levels.

### Demographic predictors of vocabulary

Education was the strongest single correlate and was not merely a proxy for reading: each survived control for the other and for age. Parental education retained a partial correlation of 0.24 once the participant’s own education was removed, reflecting the independent effect of family background. That highest household income predicted better than most recent household income (0.27 against 0.18) suggesting that scores reflect accumulated exposure to language over a lifetime rather than present circumstances. Participants who did not speak English in the home while growing up scored 5.2 levels lower (d = −0.57), a difference that persisted with education and age controlled (partial r = −0.26), and carried the largest weight in the comprehensive model. This is consistent with the bilingual lexical deficit documented by meta-analysis: bilingual speakers know fewer words in their second languages than monolingual speakers, with an effect of about half a standard deviation on vocabulary tests (Bylund et al., 2023). Marital status and family history of dementia correlated with the score only because both were age-related. There was no sex difference (d = 0.007, 95% CI −0.08 to +0.09). This is consistent with studies of other English-language vocabulary and premorbid-ability measures, including the NAART, Shipley Vocabulary, WAIS-IV Vocabulary, and the English NIH Toolbox Picture Vocabulary Test, which generally report negligible male–female differences.

Vocabulary scores were approximately 0.15 levels higher per year of age across the cross-sectional adult sample, within every education stratum, though at different rates (0.09, 0.10 and 0.18 levels per year). The magnitude agrees with large web-based estimates of vocabulary growth across adulthood: Brysbaert et al. (2016) tested 221,268 participants and estimated the gain at roughly one new lemma every two days between twenty and sixty.

### Race and demographic adjustment

We examined race/ethnicity as a social/demographic grouping variable, not a biological construct. Race/ethnic differences were among the largest seen with any CCAB test, and were attenuated, but not eliminated, with the comprehensive model: with ESL, parental education, and income in the model, the adjusted group coefficients were −0.27 to −0.38 SD for Asian, Hispanic, and Other groups, and −0.76 SD for NH Black participants, all significant. The pattern is the one described in the literature on quality of education: Manly et al. (2002) showed that reading level, a proxy for the quality rather than the quantity of schooling, attenuated Black–White differences on neuropsychological tests that years of education did not, and Manly et al. (2004) extended the finding to acculturation. Years of schooling, parental education, and income are coarse proxies for the lifetime exposure to the written and spoken lexicon that a synonym test measures. There was little evidence that the group differences arose from systematic differential item functioning: difficulties estimated separately in the NH Black, NH Asian, and Hispanic groups agreed closely with the NH White calibration (r = 0.96) across the adequately sampled portion of the bank (Table 6). Whether to add race/ethnicity indicators, which equalize group means by construction and reduce the low-score flag ratio from 8.0:1 to 2.3:1, is a scoring-policy decision rather than a statistical one.

### List ordering and the staircase

The CCAB list was ordered by aggregated play on a web game rather than by expert judgment or formal calibration. Rasch calibration showed that the ordering closely paralleled a formal calibration. Minor differences reflected usage drift: Roivainen (2014) traced Wordsum, WAIS, and WISC vocabulary items through Google Ngrams and found that usage frequency correlated .38 to .73 with item difficulty. Here the largest displacements in CCAB vocabulary items (e.g., meme, aphasia) reflected more common usage. The rank order of vocabulary items is nonetheless durable over decades (Dorius & Alwin, 2016; Malhorta et al., 2007) and the agreement of the assigned levels with the Brysbaert et al. (2019) prevalence norms (ρ = −0.87), collected from a different population using a different method, shows that the inherited ordering tracks word knowledge in English, and not only performance in this sample.

The adaptive staircase used a 1-up/1-down rule with equal steps in the two directions, which converges on the 50%-correct point (Levitt, 1971). Accuracy over the final four trials (0.55) confirmed this—with a 25% chance of a correct guess, 0.55 corresponds to knowing about 40% of the words at the final level, and the 0.50 convergence point to knowing a third— and accuracy over all 24 trials was higher (0.64) only because the sampling began with words which were below the final level of most subjects. The convergence point does not affect the QuickSIN score, which is computed from the proportion correct at every level visited rather than from where the track ended. In these data that score reproduced the model-based Rasch ability estimate at r = 0.99.

The within-level spread of difficulty has a practical implication for the resolution of level-based scores. A step of one level raised difficulty only 60% of the time, and a step of two levels 69% of the time, so the final adjustments of the staircase, which the mean of the last four averages, were close to a coin flip. The effective resolution on the level scale was therefore 2–3 levels. The Rasch ability estimate, which carries its own standard error (0.48 logits, about 2.6 levels), makes this explicit, and its higher twelve-month retest reliability (0.84 against 0.81) shows that part of the level-based score’s retest error is attributable to the ordering rather than to the examinee.

### WAIS and CCAB approaches

The WAIS-IV Vocabulary test is subjectively scored and requires 7–15 minutes for administration. It administers thirty items in a fixed sequence, starting at item 5 for standard administration and discontinuing after three consecutive scores of zero. Test length is therefore determined by performance: a strong examinee receives most of the list and a weak one stops early. This introduces dependence on local item ordering around the discontinuation point; after three consecutive zero scores, more difficult items are not sampled while in some cases an examinee who would have answered more difficult items correctly never sees them.

In the CCAB vocabulary test every participant receives 24 items; test length is fixed, and performance determines which difficulty levels are tested. The adaptive staircase settles near the participant’s threshold quickly and thus, in contrast to the WAIS approach, samples words above and below the participant’s ability level extensively. Administration time is predictable: a median of 227 s with an interquartile range of 200 to 258 s.

### Comparisons with other adaptive vocabulary tests

Two useful comparison classes are calibrated adaptive vocabulary tests and large crowdsourced vocabulary studies. The NIH Toolbox picture vocabulary test is a fully calibrated adaptive test. Its bank of 602 Rasch-calibrated words is close to the size of the CCAB stimulus set; its administered length of at most 25 items is near the CCAB 24-trial structure; and its 7-to-21-day retest intraclass correlation of .80 is close to the CCAB twelve-month ICC of 0.81 and one-day ICC of 0.83. The core difference between the two tests lies in the provenance of the word difficulties: the NIH assignments came from 4,703 participants (1,513 adults) and at least 200 responses per item under a documented Rasch calibration, whereas the CCAB assignments came from aggregated web play and were calibrated here against 2,181 adults.

Another NIH Toolbox variant, the Mobile Toolbox Word Meaning test (Young et al., 2024) more closely resembles the CCAB vocabulary test. The Word Meaning Test is a five-alternative synonym test whose 1,363-word bank was Rasch-calibrated on 7,525 online participants, retested at an intraclass correlation of 0.65 over two weeks. Because items answered correctly by fewer than 15% or more than 85% of the calibration sample were discarded, its bank has a much narrower difficulty range than CCAB vocabulary, spanning 4.3 logits against 11.2 for the CCAB bank. The adaptive rules also differ. Word Meaning begins every examinee at an ability estimate of zero, re-estimates ability after each response, and selects the next item to give a 0.675 probability of a correct answer, stopping after 15 to 25 items when the standard error falls to 0.50. The CCAB staircase begins at level 16 and adapts level rapidly in fixed steps, reducing step size after successive reversals. For an examinee who answers the first four items correctly, the CCAB staircase rises 16 levels, about 2.9 logits, whereas the Word Meaning test using an item-response estimate with a standard normal prior rises about 1 logit in decelerating steps. Thus, the CCAB staircase reaches difficult test items in fewer trials, often overshooting the participant’s final level (Figure 1). Because the Word Meaning selection rule is deterministic given the responses, examinees with the same early response pattern receive the same words unless an exposure-control procedure is applied. The CCAB test draws each word at random from the ten words at each level, so that identical level paths yield different words and a retest rarely repeats words (e.g., average two or three of the 24 words). Word Meaning reports its score as the Rasch ability estimate; the CCAB test reports a level score and, from the present calibration, is virtually identical to a Rasch ability estimate. The reported Word Meaning two-week retest intraclass correlation was 0.65 in 141 participants, against 0.83 at one day and 0.81 at twelve months for the CCAB level score and 0.84 at both intervals for the ability estimate. The demographic profiles of the CCAB and Word Meaning test are similar, both show significant correlations with age and education.

### Limitations

The sample was a volunteer telemedical sample, not a census-matched standardization sample, and its education distribution (M = 15.4 years) is above the nationwide population mean. Some demographic questions were unanswered: the parental-education, income, and social-standing items were unavailable in about 25% of participants, so the comprehensive model was fitted on 1,564 participants. Race was self-reported into four broad categories. The twenty-one demographic variables in Table 3 were not corrected for multiple comparisons, and the variables at the bottom of that table should be treated as hypotheses. The empirical-Bayes prior pulled sparsely sampled words toward the difficulty expected from their assigned levels, inflating agreement across the full bank, so the correlation among the 316 well-sampled words is the more informative estimate; 195 words at the two ends of the bank were too sparsely sampled for stable data-driven difficulty estimation; 94 words, mostly at the hard end, are absent from the prevalence norms. The invariance analyses could compare only 91–102 of the 600 words per race/ethnicity group, those answered at least 30 times in both the group and the NH White participants, and were confined to levels 16–40; their power to detect differential item functioning smaller than about 0.8 logits in a single word was low, so while they rule out a general reordering, small item-specific effects may have been missed. Because the age analyses were cross-sectional, the observed age gradient likely reflects cohort as well as within-person aging effects. Concurrent validity against an established individually administered vocabulary or premorbid-ability measure was not available in this cohort and remains to be established.

## Conclusion

The CCAB adaptive vocabulary test measured verbal ability in four minutes with a one-day retest reliability of 0.83, a twelve-month reliability of 0.81, no practice effect, and negligible floor and ceiling effects across the adult span. The vocabulary score tracked education (r = 0.43), hours reading (r = 0.39), age (r = 0.29), household income, parental education, and English-language background, but not sex. A seven-term comprehensive model explained 32% of the variance (cross-validated R^2^ = .31) against 17% for age– education–gender adjustment alone, and attenuated but did not eliminate racial/ethnic differences: adjusted differences from NH White participants were 0.27–0.38 SD for Hispanic, Asian, and other participants and 0.76 SD for Black participants. A Rasch calibration of the word bank showed that the ordering largely inherited from aggregated FreeRice response data agreed with estimated difficulty at ρ = 0.95 among well-sampled words, but that the within-level spread of about 0.53 logits limits the resolution of a level-based score to 2–3 levels. Rescoring on the calibrated scale raised twelve-month retest reliability from 0.81 to 0.84. Independent word prevalence norms agreed with the calibrated difficulties at ρ = −0.85.

## Declarations

### Ethics approval and informed consent

WIRB protocol 20201196. All participants provided written informed consent.

### Clinical trial registration

NCT04800588.

### Data availability

Data are available for reasonable requests.

### Author contributions

All authors contributed to the work and approved the final version.

### Use of artificial intelligence

Large language models (Claude, Anthropic; ChatGPT, OpenAI) were used to assist with statistical programming and generation of figure code, literature identification, and editing of the manuscript text. The authors specified the analyses, verified the code and all reported values against the data, checked every reference, and take full responsibility for the content.

